# Outcomes of Persons With COVID-19 in Hospitals With and Without Standard Treatment With (Hydroxy)chloroquine

**DOI:** 10.1101/2020.08.14.20173369

**Authors:** EJG Peters, D Collard, S Van Assen, M Beudel, MK Bomers, J Buijs, LR De Haan, W De Ruijter, RA Douma, PWG Elbers, A Goorhuis, NC Gritters van den Oever, GHH Knarren, HS Moeniralam, RLM Mostard, MJR Quanjel, AC Reidinga, R Renckens, JPW Van Den Bergh, IN Vlasveld, JJ Sikkens, On behalf of the CovidPredict Study Group

## Abstract

**Objective:** To compare survival of subjects with COVID-19 treated in hospitals that either did or did not routinely treat patients with hydroxychloroquine or chloroquine.

**Methods:** We analysed data of COVID-19 patients treated in 9 hospitals in the Netherlands. Inclusion dates ranged from February 27^th^ 2020, to May 15^th^, when the Dutch national guidelines no longer supported the use of (hydroxy)chloroquine. Seven hospitals routinely treated subjects with (hydroxy)chloroquine, two hospitals did not. Primary outcome was 21-day all-cause mortality. We performed a survival analysis using log-rank test and Cox-regression with adjustment for age, sex and covariates based on premorbid health, disease severity, and the use of steroids for adult respiratory distress syndrome, including dexamethasone.

**Results:** Among 1893 included subjects, 21-day mortality was 23.4% in 1552 subjects treated in hospitals that routinely prescribed (hydroxy)chloroquine, and 17.0% in 341 subjects that were treated in hospitals that did not. In the adjusted Cox-regression models this difference disappeared, with an adjusted hazard ratio of 1.17 (95%CI 0.88-1.55). When stratified by actually received treatment in individual subjects, the use of (hydroxy)chloroquine was associated with an increased 21-day mortality (HR 1.58; 95%CI 1.25-2.01) in the full model.

**Conclusions:** After adjustment for confounders, mortality was not significantly different in hospitals that routinely treated patients with (hydroxy)chloroquine, compared with hospitals that did not. We compared outcomes of hospital strategies rather than outcomes of individual patients to reduce the chance of indication bias. This study adds evidence against the use of (hydroxy)chloroquine in patients with COVID-19.

## Introduction

The spread of SARS-CoV-2, leading to the current pandemic of COVID-19, has a profound global impact on daily life, morbidity and mortality. Several preliminary studies have reported that the antimalarial agents hydroxychloroquine and chloroquine, or (H)CQ, alone or in combination with the antibiotic azithromycin, can have a suppressive effect on the viral replication, and might decrease the mortality of COVID-19^1-4^. So far, clinical studies have been hampered by (indication) bias^1,2,4^, monocentre setup^2,3^, small numbers of included subjects^3^, and concerns about the verifiability of data, even leading to withdrawal of a publication ^4^. Side effects of (H)CQ are well-known and common, and include fever and cardiac arrhythmias. Randomized controlled clinical trials (RCTs) are currently being conducted to investigate the effect of (H)CQ on outcome of COVID-19. While we are awaiting definite results from RCTs, cohort studies can provide quick closure of existing knowledge gaps. When treatment assignment in cohort studies is based on prescriber discretion, the risk of indication bias (even after covariate adjustment) remains high. However, our database of Dutch hospitals contains data of subjects from hospitals that either routinely prescribed (H)CQ or did not prescribe it at all, offering a unique opportunity to compare both strategies. The comparison of different treatment strategies among hospitals leads to a significant reduction of (indication) bias. The objective of this study was to compare the effect of hospital-wide COVID-19 treatment strategies with or without routine (H)CQ use on all-cause 21-day mortality.

## Methods

We used data from the ongoing CovidPredict Clinical Course Cohort containing over 2,000 persons with COVID-19^5^, from 9 hospitals in the Netherlands, including two tertiary care hospitals. Included in the database were all subjects admitted to hospital with positive SARS-CoV-2 PCR of nasopharynx, throat, sputum or bronchoalveolar lavage samples, or CT-scan abnormalities that were typical for COVID-19 (CO-RADS 4 and 5)^6^, without another explanation for the abnormalities than COVID-19. Inclusion dates ranged from the first admitted case in the Netherlands on February 27^th^ 2020, to May 15^th^, when the Dutch national guidelines no longer advised the use of (H)CQ. We excluded patients < 18 years and patients who were transferred to or from another hospital. Dosage of chloroquine base was: loading dose of 600 mg, followed by 300 mg twice a day for a total of 5 days. Dosage of hydroxychloroquine sulphate was 400 mg twice daily on the first day, followed by 200 mg twice daily on days 2 to 5. Among the seven (H)CQ-hospitals, the timing of start of (H)CQ treatment differed; three hospitals started at the moment of COVID-19 diagnosis, four started after diagnosis but only when patients clinically deteriorated e.g., when there was an increase in respiratory rate or increase in use of supplemental oxygen. The two hospitals that did not routinely treat subjects with (H)CQ (i.e., the non-(H)CQ-hospitals), offered best supportive care, including oxygen therapy and potentially antibiotic therapy, according to local guidelines and prescriber discretion. Participating hospitals did not routinely prescribe other experimental medication (e.g., lopinavir/ritonavir, remdesivir or steroids, see Table 1). Subjects who were incidentally treated with these drugs were included in the study. Primary outcome was 21-day all-cause mortality, defined as hospital mortality, or discharge to a hospice care facility. A waiver for the use of hospital record data was obtained through the Institutional Review Board of Amsterdam UMC; however, patients were given the opportunity to opt out. In the primary analysis, we compared effectiveness of (H)CQ versus non-(H)CQ hospital strategies, irrespective of actual individual (H)CQ treatment. We performed a survival analysis using log-rank test and Cox-regression with adjustment for age, sex and covariates based on premorbid health (i.e., history of lung, kidney and cardiovascular disease, diabetes mellitus, obesity, and neoplasms or hematologic disease), disease severity during presentation (respiratory rate, oxygen saturation) and the use of steroids, including dexamethasone, for adult respiratory distress syndrome (ARDS)^7,8^. We collected data according to the collection protocol of the World Health Organization. Missing covariates were imputed using multiple imputation. As a sensitivity analysis, we performed a complete case analysis using inverse probability weighting of propensity scores (determined using the same covariates), and a subgroup analysis in hospitals who started (H)CQ from the moment of diagnosis. Finally, we repeated the analyses comparing actually received treatment, with (H)CQ. All statistical analyses were performed using R versions 3.6.3 (R Foundation, Vienna, Austria).

**Table 1:**
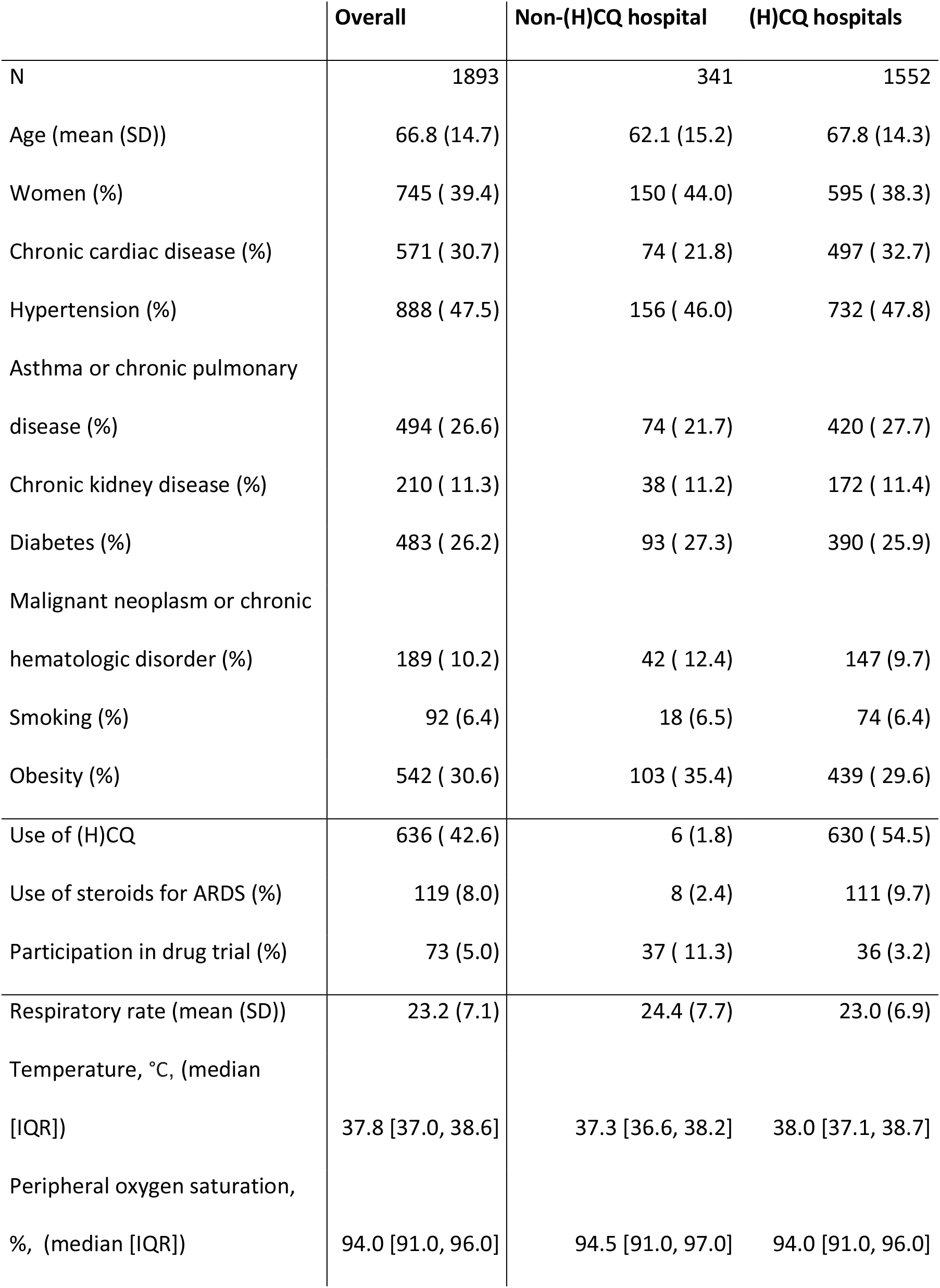
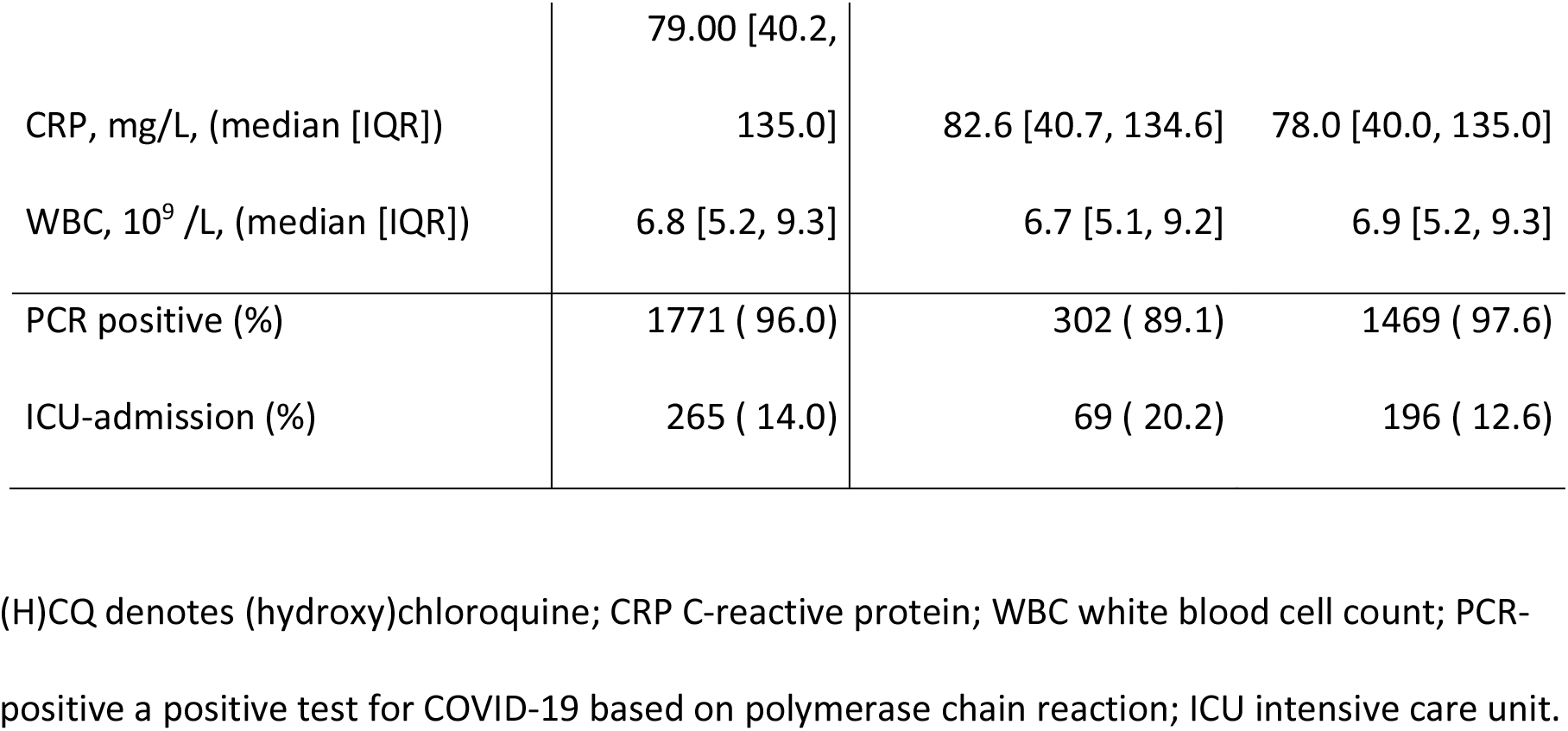
Baseline characteristics.

## Results

We analysed results of 1893 subjects admitted before May 15^th^ 2020, 239 were excluded because they were transferred from another hospital. Demographic data are shown in Table 1. Follow-up data were missing for 28 (2.5%) subjects. In total, 1552 subjects were treated in hospitals where (H)CQ was a standard part of treatment strategy ((H)CQ hospitals) and 341 in hospitals where (H)CQ was not a standard part of treatment (non-(H)CQ hospitals). In (H)CQ-hospitals, 54.5% of the subjects received (H)CQ, compared with 1.8% of the subjects in the non-(H)CQ-hospitals. Among the seven (H)CQ-hospitals, two used hydroxychloroquine during the first half and chloroquine during the second half of the epidemic, whereas five hospitals used chloroquine only. Subjects in (H)CQ-hospitals were older (68 (SD: 14) vs 62 (SD: 15) years) and had a higher prevalence of chronic pulmonary disease (27.7 vs 21.7%) than subjects in the non-(H)CQ-hospitals. Respiratory rate and peripheral oxygen saturation during admission were similar in both hospital groups (see Table 1). In (H)CQ-hospitals, 9.7% of subjects received corticosteroids for ARDS and 3.2% were in a study protocol of an experimental SARS-CoV-2 directed antiviral (e.g., lopinavir/ritonavir) or immunomodulatory drug trial (e.g., imatinib, anti-complement C5), versus 2.4% and 11.3% in non-(H)CQ-hospitals, respectively. Figure 1 shows the survival of subjects in (H)CQ-versus non-(H)CQ-hospitals. Unadjusted mortality at day 21 was significantly different between the (H)CQ- and non-(H)CQ-hospitals (23.4% vs. 17.0%). However, in the adjusted Cox-regression models, this difference disappeared, with an adjusted hazard ratio of 1.17 (95%CI 0.88-1.55, Table 2). The sensitivity analysis of hospitals routinely starting (H)CQ treatment from the moment of COVID-19 diagnosis (i.e., (H)CQ hospitals without the hospitals that initiated (H)CQ treatment upon clinical deterioration) compared with non-(H)CQ-hospitals, showed similar results with a HR of 1.14 (95%CI 0.82-1.57) after adjustment for age, sex, comorbidities, and disease severity at presentation. Complete case analysis and the analysis using propensity scores showed similar results (see Table 3). Finally, when stratified by actually received treatment, the use of (H)CQ was associated with an increased 21-day mortality (HR 1.58; 95%CI 1.25-2.01, Table 3) in the full model. The sensitivity analysis of all hospitals and all subjects can be found in Table 4.

**Figure 1:**
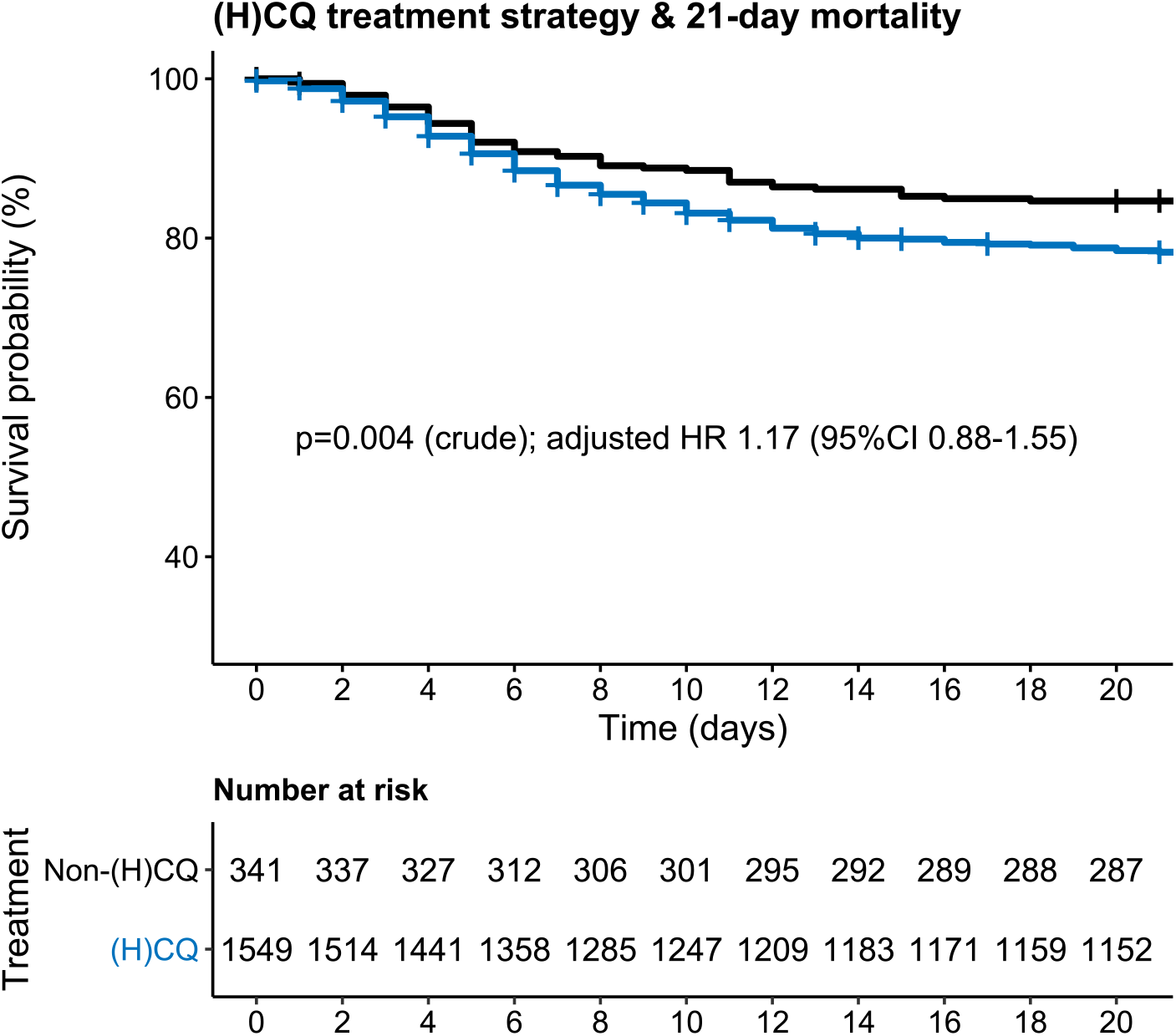
Mortality of subjects in the (H)CQ-hospitals (blue) versus non-(H)CQ-hospitals (black).

**Table 2:**
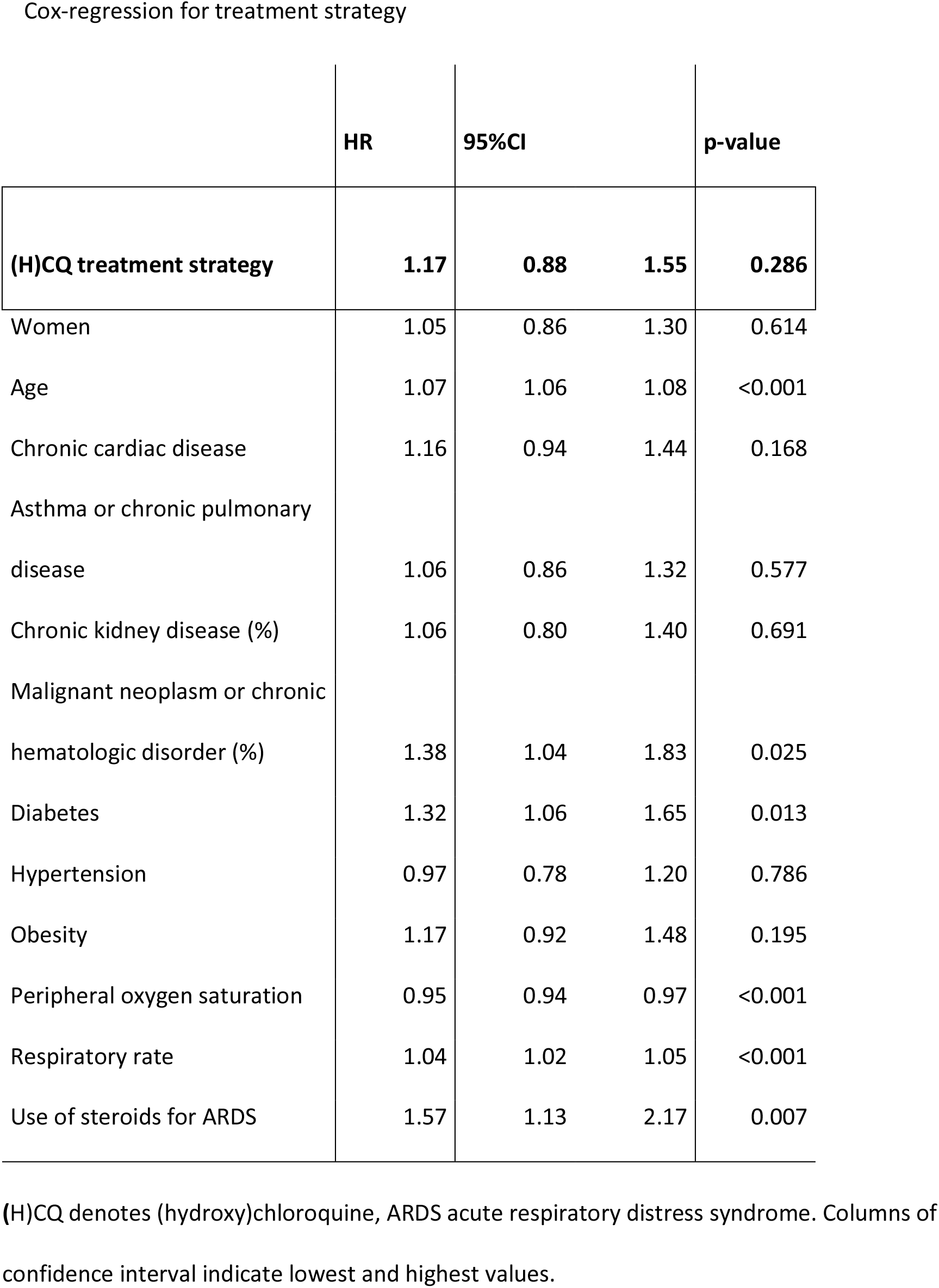
Results of Cox-regression models after stratification for treatment strategy.

|  | HR | 95%CI |  | p-value |
| --- | --- | --- | --- | --- |
| **(H)CQ treatment strategy** | **1.17** | **0.88** | **1.55** | **0.286** |
| Women | 1.05 | 0.86 | 1.30 | 0.614 |
| Age | 1.07 | 1.06 | 1.08 | <0.001 |
| Chronic cardiac disease | 1.16 | 0.94 | 1.44 | 0.168 |
| Asthma or chronic pulmonary disease | 1.06 | 0.86 | 1.32 | 0.577 |
| Chronic kidney disease (%) | 1.06 | 0.80 | 1.40 | 0.691 |
| Malignant neoplasm or chronic hematologic disorder (%) | 1.38 | 1.04 | 1.83 | 0.025 |
| Diabetes | 1.32 | 1.06 | 1.65 | 0.013 |
| Hypertension | 0.97 | 0.78 | 1.20 | 0.786 |
| Obesity | 1.17 | 0.92 | 1.48 | 0.195 |
| Peripheral oxygen saturation | 0.95 | 0.94 | 0.97 | <0.001 |
| Respiratory rate | 1.04 | 1.02 | 1.05 | <0.001 |
| Use of steroids for ARDS | 1.57 | 1.13 | 2.17 | 0.007 |

(H)CQ denotes (hydroxy)chloroquine, ARDS acute respiratory distress syndrome. Columns of confidence interval indicate lowest and highest values.

**Table 3:** Results of Cox-regression models after stratification for actual treatment.

|  | HR | 95%CI |  | p-value |
| --- | --- | --- | --- | --- |
| **(H)CQ treatment** | 1.58 | 1.25 | 2.01 | <0.001 |
| Women | 1.09 | 0.88 | 1.34 | 0.439 |
| Age | 1.07 | 1.06 | 1.08 | <0.001 |
| Chronic cardiac disease | 1.21 | 0.97 | 1.50 | 0.092 |
| Asthma or chronic pulmonary<br>disease | 1.05 | 0.85 | 1.30 | 0.662 |
| Chronic kidney disease (%) | 1.07 | 0.81 | 1.42 | 0.621 |
| Malignant neoplasm or chronic<br>hematologic disorder (%) | 1.41 | 1.06 | 1.87 | 0.018 |
| Diabetes | 1.30 | 1.05 | 1.63 | 0.018 |
| Hypertension | 0.97 | 0.78 | 1.20 | 0.777 |
| Obesity | 1.17 | 0.92 | 1.48 | 0.191 |
| Peripheral oxygen saturation | 0.96 | 0.94 | 0.97 | <0.001 |
| Respiratory rate | 1.03 | 1.02 | 1.05 | <0.001 |
| Use of steroids for ARDS | 1.45 | 1.03 | 2.05 | 0.033 |

(H)CQ denotes (hydroxy)chloroquine, ARDS acute respiratory distress syndrome.

**Table 4:** Sensitivity Analysis.

|  | HR | 95%CI | p-value |
| --- | --- | --- | --- |
| **(H)CQ received treatment** | 1.48 | 1.26 1.73 | <0.001 |

Complete case analysis using inverse probability weighting For **treatment strategy**
|  | HR | 95%CI | p-value |
| --- | --- | --- | --- |
| **(H)CQ treatment strategy** | 1.21 | 1.03 1.44 | 0.024 |

(H)CQ denotes (hydroxy)chloroquine.

## Discussion

At first glance, our study results may seem to suggest that subjects treated in hospitals that routinely prescribed (H)CQ had a significantly increased 21-day all-cause mortality compared with those in hospitals that did not routinely prescribe (H)CQ. However, mortality was not significantly different after adjustment for age, sex, medical history, disease severity at presentation and steroid use during treatment. Similarly, we found an increased risk of death among subjects who had actually received treatment with (H)CQ, which has likely been driven by indication bias, as in four of the seven (H)CQ-hospitals, (H)CQ was only prescribed upon clinical deterioration. The unique characteristics of our study cohort enabled a study design that minimized indication bias. Our results add further weight to existing evidence against the use of (H)CQ for the treatment COVID-19.

The strength of this study is that data were collected in nine secondary and tertiary care hospitals in the Netherlands during the COVID-19 epidemic. Data collection was set up prospectively and the database included data on all consecutive subjects admitted to general medicine and pulmonology wards, and to intensive care units. The database was set up according to the WHO standards, which enabled data comparison and uniformity of data among the different participating centres. The comparison of hospital-defined treatment strategies rather than the treatment actually received led to a lower risk of indication bias compared with previous studies^1,2^. We roughly estimate the extend of the effect of indication bias to be the difference in outcome between the uncorrected and the corrected model. Further strengths include the multicentre setup^2,3^, as mentioned above, and the relatively large numbers of included subjects^3^.

There are some limitations we need to address. Although health care in the Netherlands has a homogeneous setup, there was some variability in standard protocols among the hospitals that could have led to residual confounding. The two non-(H)CQ-hospitals were tertiary (academic) centres, whereas the (H)CQ-hospitals comprised secondary care hospitals. Since we excluded subjects transferred to and from other hospitals, the referral role of the tertiary care hospitals was minimized. Furthermore, subjects in the (H)CQ hospitals were more likely to receive steroid treatment, while subjects in the non-(H)CQ hospitals were more likely to receive other experimental immunomodulatory drugs. The numbers of the individual types of medication were small, making it impossible to draw conclusions from these differences. The results of the RECOVERY trial (publication pending), suggested a lower mortality in patients treated with dexamethasone. Treatment with dexamethasone could therefore have resulted in a lower mortality in the group of (H)CQ hospitals. We did not find such an effect, even after correction in the full model. We also used extensive covariate adjustments, using various methods to minimize influence of differences in patient population among hospitals, and the similarity in outcomes between these methods is reassuring in this regard. Finally, because not every subject in the (H)CQ-hospitals actually received (H)CQ, the current efficacy estimate in our study is likely an underestimation of the true (H)CQ effect. Performing an instrumental variable analysis would have provided an approximation of this true effect, but because the current efficacy point estimates point toward harm rather than benefit of (H)CQ, this likely would not have changed our conclusions.^9^

Despite the positive results of some studies resulting in widespread use of (H)CQ, our study did not show a benefit of (H)CQ treatment. This may be explained by the timing of the administration of the drug and its specific working mechanism. Chloroquine binds *in silico* and *in vitro* with high affinity to sialic acids and gangliosides of SARS-CoV-2. These bindings inhibit the interaction at non-toxic plasma levels with ACE-2 receptors and could hypothetically stop the cascade from formation of pulmonary infiltrations to full blown ARDS and death^10-12^. The antiviral activity might be more effective in the pre-clinical setting as the deterioration in the hospital is more an effect of the cytokine storm provoked by SARS-CoV-2 than an effect of the viral infection itself. This hypothesis might explain why the clinical benefit for admitted subjects was absent in our study, although we did not observe a difference in outcome among subjects treated early (at diagnosis) and among those treated later upon clinical deterioration. Currently, clinical trials are underway to study (hydroxy)chloroquine in admitted COVID-19 patients (e.g., NCT04261517 and NCT04307693). As post-exposure prophylaxis, hydroxychloroquine does not seem to be effective to prevent COVID-19,^13^ but results of further studies are pending^14^. Given the current evidence, we would argue against the use (H)CQ outside the setting of randomized clinical trials.

## Data Availability

The data that support the findings of this study are available from the corresponding author, upon reasonable request.

## Acknowledgements

We would like to acknowledge the contribution of the CovidPredict Study Group for the data collection, and dr. B.J.H. van den Born and prof. dr. A.H. Zwinderman for their help with the analysis and interpretation of the data.

## Contribution of authors

All authors have made substantial contributions to the following: (1) the conception and design of the study, or acquisition of data, or analysis and interpretation of data, (2) drafting the article or revising it critically for important intellectual content, (3) final approval of the version to be submitted.

## References

1. Rosenberg ES, Dufort EM, Udo T, et al. Association of Treatment With Hydroxychloroquine or Azithromycin With In-Hospital Mortality in Patients With COVID-19 in New York State. JAMA 2020.

2. Geleris J, Sun Y, Platt J, et al. Observational Study of Hydroxychloroquine in Hospitalized Patients with Covid-19. N Engl J Med 2020.

3. Gautret P, Lagier JC, Parola P, et al. Hydroxychloroquine and azithromycin as a treatment of COVID-19: results of an open-label non-randomized clinical trial. Int J Antimicrob Agents 2020:105949.

4. Mehra MR, Desai SS, Ruschitzka F, Patel AN. Hydroxychloroquine or chloroquine with or without a macrolide for treatment of COVID-19: a multinational registry analysis. Lancet 2020.

5. https://covidpredict.nl/en. 2020. (Accessed 9 June, 2020,

6. Prokop M, van Everdingen W, van Rees Vellinga T, et al. CO-RADS - A categorical CT assessment scheme for patients with suspected COVID-19: definition and evaluation. Radiology 2020:201473.

7. Yang X, Yu Y, Xu J, et al. Clinical course and outcomes of critically ill patients with SARS-CoV-2 pneumonia in Wuhan, China: a single-centered, retrospective, observational study. Lancet Respir Med 2020;8:475–81.

8. Petrilli CM, Jones SA, Yang J, et al. Factors associated with hospital admission and critical illness among 5279 people with coronavirus disease 2019 in New York City: prospective cohort study. BMJ 2020;369:m1966.

9. Instrumental variable analysis. 2020, at https://journals.lww.com/epidem/Fulltext/2006/05000/Evaluating_Short_Term_Drug_Effects_Using_a.11.aspx.)

10. Fantini J, Di Scala C, Chahinian H, Yahi N. Structural and molecular modelling studies reveal a new mechanism of action of chloroquine and hydroxychloroquine against SARS-CoV-2 infection. Int J Antimicrob Agents 2020;55:105960.

11. Fan J, Zhang X, Liu J, et al. Connecting hydroxychloroquine in vitro antiviral activity to in vivo concentration for prediction of antiviral effect: a critical step in treating COVID-19 patients. Clin Infect Dis 2020.

12. Yao X, Ye F, Zhang M, et al. In Vitro Antiviral Activity and Projection of Optimized Dosing Design of Hydroxychloroquine for the Treatment of Severe Acute Respiratory Syndrome Coronavirus 2 (SARS-CoV-2). Clin Infect Dis 2020.

13. Boulware DR, Pullen MF, Bangdiwala AS, et al. A Randomized Trial of Hydroxychloroquine as Postexposure Prophylaxis for Covid-19. N Engl J Med 2020.

14. Mitja O, Clotet B. Use of antiviral drugs to reduce COVID-19 transmission. Lancet Glob Health 2020;8:e639-e40.

